# Rapid processing of SARS-CoV-2 containing specimens for direct RT-PCR

**DOI:** 10.1101/2020.12.14.20248183

**Authors:** Piotr Chomczynski, Peter W. Chomczynski, Amy Kennedy, Michal Rymaszewski, William W. Wilfinger, Judith A. Heiny, Karol Mackey

## Abstract

Widespread diagnostic testing is needed to reduce transmission of COVID-19 and manage the pandemic. Effective mass screening requires robust and sensitive tests that reliably detect the SARS-CoV-2 virus, including asymptomatic and pre-symptomatic infections with a low viral count. Currently, the most accurate tests are based on detection of viral RNA by RT-PCR. We developed a method to process COVID-19 specimens that simplifies and increases the sensitivity of viral RNA detection by direct RT-qPCR, performed without RNA purification. In the method, termed Alkaline-Glycol Processing (AG processing), a SARS-CoV-2-containing biological specimen, such as saliva or a swab-collected suspension, is processed at pH 12.2 to 12.8 for 5 min at room temperature. An aliquot of the AG-processed specimen is used for detection of SARS-CoV-2 RNA by direct RT-qPCR. AG processing effectively lyses viruses and reduces the effect of inhibitors of RT-PCR that are present in biological specimens. The sensitivity of detecting viral RNA using AG processing is on par with methods that include a viral RNA purification step. One copy of SARS-CoV-2 virus per reaction, equivalent to 300 copies per ml of saliva, is detectable in the AG-processed saliva. The LOD calculated following U.S. FDA guidelines is 600 viral copies per ml of initial saliva specimen. AG processing works with saliva specimens or swab specimens collected into Universal Transport Medium (UTM), is compatible with heat treatment, and was confirmed to work with a range of CDC-approved RT-qPCR products and kits. Detection of SARS-CoV-2 RNA using AG processing with direct RT-qPCR provides a reliable and scalable diagnostic test for COVID-19 that can be integrated into a range of workflows, including automated settings.

## Introduction

The COVID-19 pandemic has created a great demand for molecular tests that detect viral infection by severe acute respiratory syndrome coronavirus 2 (SARS-CoV-2), the RNA virus responsible for coronavirus disease 2019 (COVID-19). Currently, the most accurate diagnostic tests for COVID-19 are based on detection of SARS-CoV-2 RNA. However, virus-containing biological specimens collected from patients contain components that negatively affect detection of viral RNA by degrading RNA and inhibiting RT-PCR. Several strategies have been developed to mitigate the negative actions of these components.

Commonly used tests to detect viral RNA involve removal of extraneous components from a biological specimen by RNA purification. These tests include two steps: 1) a purification step to extract viral RNA from a patient sample (nasopharyngeal swab, sputum, or other); followed by 2) a reverse transcription-quantitative PCR (RT-qPCR) assay to detect SARS-CoV-2 RNA. Effective target sequences for detecting SARS-CoV-2 RNA by RT-qPCR are located in the nucleocapsid (N) gene or the spike (S) protein gene of SARS-CoV-2, and the most effective target is the N1 sequence of the N gene [1]. The lower limit of detection (LOD) of RT-qPCR performed on samples of purified SARS-CoV-2 RNA using N1 primers is approximately 10 copies of viral RNA per reaction, with a cycle threshold (C_T_) of 36 [2]. For automated detection of SARS-CoV-2 using cartridges for viral RNA isolation followed by RT-PCR, the LOD for detecting viral RNA was 14.8 copies per reaction and 365 copies per ml of a viral specimen, using primers for the S gene [3]. Alternatively, RT and a Loop-Mediated Isothermal Amplification (LAMP) method can be used to detect SARS-CoV-2 RNA. The detection sensitivity of an RT-LAMP test for SARS-CoV-2 was 100 copies of purified viral RNA per reaction [4].

Other RT-qPCR tests have been developed that eliminate the viral RNA purification step, in which a minimally processed virus-containing specimen is added directly to the RT-qPCR assay (direct RT-qPCR). Direct RT-qPCR offers advantages for streamlining testing because it is faster and simpler to implement than tests which require an initial RNA purification step. Most direct RT-PCR tests use saliva because it is more sensitive than nasopharyngeal swabs for detection of SARS-CoV-2 and it is simpler and safer to collect [5]. Direct RT-qPCR diagnostic tests have been developed in which heat-processed saliva samples or viral suspensions in Universal Transport Medium (UTM) are diluted in a buffer, and an aliquot of the treated sample is added directly to a reagent mix to perform RT-PCR, without RNA purification. A direct RT-qPCR test with a viral suspension heated at 95 C for 10 min has been described [6]. For a low viral load (C_T_ > 30), this method was not satisfactory and test results required further confirmation. In other direct RT-PCR tests, saliva was diluted with a buffer and heated at 95 C for 30 min, then supplemented with Tween-20 [7], or heated at 65 C for 10 min [8]; an aliquot of the heat-treated, diluted sample was used for direct RT-PCR. The LOD for these tests, evaluated using saliva spiked with γ-irradiated SARS-CoV-2, was 1,000 copies of SARS-CoV-2 virus per ml of saliva [7], or 6,600 per ml of a viral suspension spiked with synthetic SARS-CoV-2 RNA [8]. Another direct RT-PCR test for detecting SARS-CoV-2 RNA used saliva treated with dithiothreitol (Sputasol) and a ribonuclease inhibitor [9]. For this method, the LOD of the final RT-PCR mix, evaluated using saliva spiked with a viral RNA template of 99 bp, was 12 copies of RNA template per reaction (equivalent to 14,000 copies of viral RNA template per ml of saliva). Another publication described a method in which saliva samples were treated with proteinase K and then heated before being used in a direct RT-PCR assay [10]. The reported LOD for detecting SARS-CoV-2, evaluated using SARS-CoV-2 positive saliva, was 6,000 virus copies per ml of saliva. In direct RT-PCR tests, addition of non-ionic detergents, such as Tween-20 and Triton X-100, to saliva or swab-derived samples was shown to increase the sensitivity of SARS-CoV-2 RNA detection [7,11].

Our goal was to further simplify detection of SARS-CoV-2 RNA by direct RT-PCR and to enhance its sensitivity. We focused on the initial processing of clinical specimens because this step is time-consuming and critically affects the sensitivity of subsequent RNA detection assays. We developed a method, termed Alkaline-Glycol Processing (AG processing), patent pending [12], in which a virus-containing specimen is briefly incubated in an alkaline-glycol solution and the AG-processed specimen is assayed for the presence of a viral RNA by direct RT-qPCR. The method builds on a previously described direct PCR assay for detecting DNA in biological samples [13,14]. It was found that concentrated polyglycols in alkaline aqueous solution act as chaotropes, which increase the alkalinity of a lysis solution and promote alkaline lysis of biological samples.

We tested detection of viral RNA using AG processing with human saliva specimens from multiple donors and viral-containing specimens suspended in Universal Transport Media (UTM) or Hanks’ medium. Direct RT-qPCR assays were performed with a range of RT-qPCR products and kits, using γ-irradiated or heat-inactivated virus particles as template. We determined the LOD for detecting viral RNA using saliva spiked with a defined quantity of γ-irradiated or heat-inactivated SARS-CoV-2 virus particles or a defined quantity of synthetic SARS-CoV-2 viral RNA. We report that AG processing used with direct RT-qPCR provides a highly accurate and sensitive diagnostic test for COVID-19. The method is economical, scalable, and can be integrated into a range of diagnostic workflows and automated settings.

## Methods

### RT-qPCR

RT-qPCR tests were performed using commercially-available one-step RT-qPCR kits. The following kits were used: TaqPath™ 1-Step RT-qPCR Master Mix, CG (Thermo Fisher Scientific, Waltham, MA, USA; cat. no. A15300). qScript™ XLT 1-Step RT-qPCR ToughMix™ ROX™ (Quantabio, Beverly, MA, USA ; cat. no. 95133-500). GoTaq® Probe 1-Step RT-qPCR System (Promega Corporation, Madison WI, USA; cat. no A6121). KiCqStart® Probe qPCR ReadyMix™, ROX™ (MilliporeSigma, St. Louis, MO, USA; cat. no. KCQS06). The RT-qPCR reaction parameters followed each manufacturer’s recommendations.

### Reference materials

The following materials were used as substitutes for SARS-CoV-2 live virus: NIAID, NIH:SARS-Related Coronavirus 2, Isolate USA-WA1/2020, Heat inactivated (NR-52286; 1.16 x 10^9^ genome equivalents per ml, BEI Resources, Manassas, VA, USA); γ-irradiated SARS-CoV-2 (NR-52287, 1.7 x 10^9^ genome equivalents per ml, BEI Resources, Manassas, VA, USA); synthetic SARS-CoV-2 RNA, 99.9% genome coverage, Control 1 (MT007544.1) and Control 2 (MN 908947.3) (Twist Bioscience, San Francisco, CA, USA). Positive control plasmid contained SARS-CoV-2 nucleocapsid gene cDNA at 2 x 10^5^ copies/µl (2019-nCoV_N Positive Control Plasmid, Integrated DNA Technologies, Coralville, IA, USA. Cat no. 10006625). Primers and detection probes for N1 and N2 regions of the nucleocapsid gene were sourced from Integrated DNA Technologies (cat. no. 10006770) that were manufactured using the U.S. CDC sequences and QC qualified under a U.S. CDC Emergency Use Authorization. The human RPP30 gene was used to confirm amplification of a gene known to be expressed in saliva (RPP30 primer set, exon 1-2, Integrated DNA Technologies, Coralville, IA, USA, Cat. no. Hs.PT.58.19785851).

### Human saliva specimens

Human saliva samples (sputum) from individual donors were collected by Molecular Research Center, Inc, in accordance with an approved IRB protocol (MRC C19 protocol, “Collection of saliva for detection of viral ribonucleic acid (RNA)”, IntegReview IRB, Austin, TX.)

### Alkaline-glycol processing of saliva

Saliva was spiked with known quantities of a SARS-CoV-2 reference material. AG processing was performed by mixing one volume of an aqueous solution containing 65% (v/v) polyethylene glycol 200 (PEG200, CAS 25322-68-3) and 45 mM KOH, with two volumes of saliva or with two volumes of universal transport medium or Hanks ‘medium containing saliva. The resulting AG processing composition contained: 21.7% (v/v) PEG200, 67 % (v/v) saliva, and 15 mM KOH, with pH ranging from 12.2 to 12.8 (depending pH of saliva specimens). AG processing was performed by incubating samples at room temperature for 5 to 30 min.

### Direct RT-qPCR detection of SARS-CoV-2 RNA

An aliquot of 5 µl of the AG processed composition was added to 15 µl of RT-qPCR reaction mix; thus, each RT-qPCR reaction contained 3.3 µl of processed saliva. RT-qPCR was performed for 45 cycles (fast cycling) following the protocol specified for each manufacturer’s kit. Reaction mixes were prepared using master mix, qPCR primer-probe mix, and Tris-HCl at pH 8 or 8.5, in accordance with the optimal conditions specified by each manufacturer. Where specified, the reaction mix included 0.2 % Tween-20. Where indicated, some saliva samples were heated at 65 or 95 C for 30 minutes prior to AG processing. Each reaction was performed in triplicate and results are presented as the mean C_T_ value. Each reaction plate included control reactions: positive control reactions consisting of synthetic SARS-CoV-2 RNA or SARS-CoV2 cDNA, processing composition reactions with no reference material, and reactions with no processing composition nor reference material. Results were considered valid if all controls provided positive or negative results, respectively.

## Results and Discussion

The RNA genome of SARS-CoV-2, like other RNA viruses, is protected by a protein-lipid envelope [15,16]. This envelope must be lysed to release RNA and make RNA available as template in molecular assays. A biological sample such as saliva contains carbohydrate and protein components, including ribonucleases that degrade RNA and inhibit RT-PCR. The alkaline-glycol processing method developed in this study lyses virus particles and suppresses RT-PCR inhibitors. These combined actions allow for highly sensitive detection of viral RNA in saliva by direct RT-qPCR, without RNA purification. The high alkaline pH in AG processing was maintained by both a strong mineral base and glycols. In this report, AG processing was performed using a solution containing (v/v): 67% of saliva, 21.6% of polyethylene glycol 200 and 15-mM KOH. Depending on the initial pH of a saliva specimen, the pH during AG processing ranged from 12.2 to 12.8.

**Table 1** compares C_T_ values for detecting SARS-CoV-2 RNA in human saliva by direct RT-qPCR performed with or without AG processing. Saliva samples were spiked with 10,000 copies of γ-irradiated or heat irradiated SARS-CoV-2 virus, or synthetic SARS-CoV-2 RNA. The N1 and N2 primer and probe sets amplified and identified target sequences in the nucleocapsid region of SARS-CoV-2 RNA. Both N1 and N2 primers amplified target sequences to a comparable degree, with the N1 primers performing marginally better. In compositions containing γ-irradiated or heat-inactivated virus, AG processing decreased the C_T_ for detecting SARS-CoV-2 RNA by about 5 amplification cycles, indicating a 32-fold increase in detection sensitivity. These results document detection of SARS-CoV-2 RNA in AG-processed human saliva and show that AG processing significantly increases the detection sensitivity for SARS-CoV-2 RNA.

**Table 1.** Detection of SARS-CoV-2 RNA in AG processed saliva by RT-qPCR.

| Template material | Processing Composition |  | N1 target |  | N2 target |  |
| --- | --- | --- | --- | --- | --- | --- |
|  |  |  | C<sub>T</sub> | ΔC<sub>T</sub> | C<sub>T</sub> | ΔC<sub>T</sub> |
| γ-irradiated SARS-CoV-2 | Saliva | AG solution | 24.16 | 5.91 | 25.09 | 5.82 |
|  |  | Water | 30.07 |  | 30.91 |  |
|  | Water | AG solution | 23.28 | 5.96 | 24.24 | 5.64 |
|  |  | Water | 29.24 |  | 29.88 |  |
| heat-inactivated SARS-CoV-2 | Saliva | AG solution | 25.07 | 4.68 | 26.90 | 4.90 |
|  |  | Water | 29.75 |  | 31.80 |  |
|  | Water | AG solution | 27.34 | 9.45 | 29.46 | 8.99 |
|  |  | Water | 36.79 |  | 38.45 |  |
| SARS-CoV-2 synthetic RNA | Saliva | AG solution | 27.15 | N/A | 28.18 | 11.80 |
|  |  | Water | Undetected |  | 39.98 |  |
|  | Water | AG solution | 26.33 | -0.13 | 27.41 | 0.11 |
|  |  | Water | 26.20 |  | 27.52 |  |

AG processing of saliva and direct RT-qPCR were performed as described in Methods. AG processing compositions contained one volume of alkaline glycol solution (AG solution) plus two volumes of saliva or water. Saliva control processing compositions contained water, saliva and no alkaline glycol solution. Water control compositions contained water and no alkaline glycol solution nor saliva. Saliva was spiked with $1.00 \times 10^4$ copies of the indicated template material per RT-qPCR reaction. RT-qPCR was performed using Sigma KiCqStart One-Step Probe RT-qPCR ReadyMix ROX using reaction parameters following the manufacturer's protocol. C<sub>T</sub> values report the mean of 3 replicates.

An even greater effect of AG processing was observed in tests using synthetic SARS-CoV-2 RNA as template. In AG-processed saliva, the presence of synthetic viral RNA was detected at C_T_27, whereas without AG processing, viral RNA was undetectable using N1 primers and barely detectable using N2 primers. This result indicated that SARS-CoV-2 RNA in untreated saliva was rapidly degraded by saliva ribonucleases, rendering it unavailable as template for RT-qPCR. Collectively, the results in Table 1 suggest that AG processing increases the detection sensitivity for viral RNA by dual actions: it promotes release of RNA from the virus capsid, providing more usable RNA template for RT-qPCR, and it protects the RT-qPCR assay from inhibitory components in saliva.

It is notable that detection of viral RNA was not impaired by the high alkaline pH of AG processing (typically pH 12.2 to pH 12.8), which is known to fragment RNA. A likely explanation for this finding is that limited fragmentation of the large SARS-CoV-2 RNA genome during AG processing did not preclude effective detection of short RNA fragments. Most RT-qPCR products are less than 100 bp in length. Thus, limited fragmentation does not preclude detection of short amplicons such as the 71 bp N1 and 67 bp N2 sequences in SARS-CoV-2.

As evidenced in **Table 2**, 5 minutes of AG processing was sufficient for effective detection of SARS-CoV-2 RNA. Extending AG processing time to 1 hour did not alter detection.

**Table 2.**
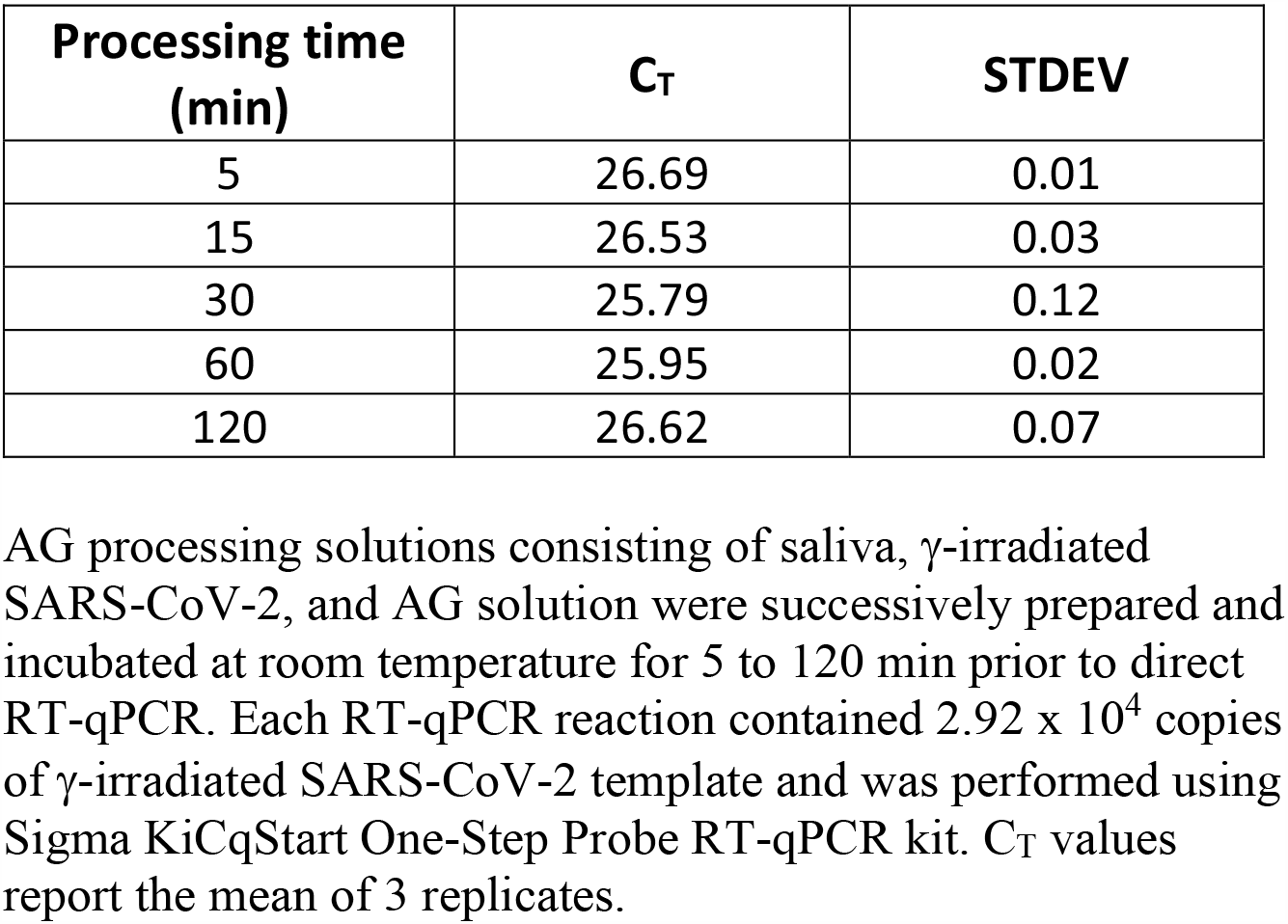
Effect of extending AG processing time on C_T_ values obtained by direct RT-qPCR.

| Processing time (min) | C<sub>T</sub> | STDEV |
| --- | --- | --- |
| 5 | 26.69 | 0.01 |
| 15 | 26.53 | 0.03 |
| 30 | 25.79 | 0.12 |
| 60 | 25.95 | 0.02 |
| 120 | 26.62 | 0.07 |

AG processing solutions consisting of saliva, γ-irradiated SARS-CoV-2, and AG solution were successively prepared and incubated at room temperature for 5 to 120 min prior to direct RT-qPCR. Each RT-qPCR reaction contained $2.92 \times 10^4$ copies of γ-irradiated SARS-CoV-2 template and was performed using Sigma KiCqStart One-Step Probe RT-qPCR kit. C<sub>T</sub> values report the mean of 3 replicates.

In addition to the Sigma KiCqStart kit used in Tables 1 & 2, AG processing used with other commercially available RT-qPCR kits likewise improved detection of SARS-CoV-2 RNA (**Table 3**). The increase in sensitivity was 32-to 70-fold, as measured by a decrease in the C_T_ needed to detect viral RNA.

**Table 3.**
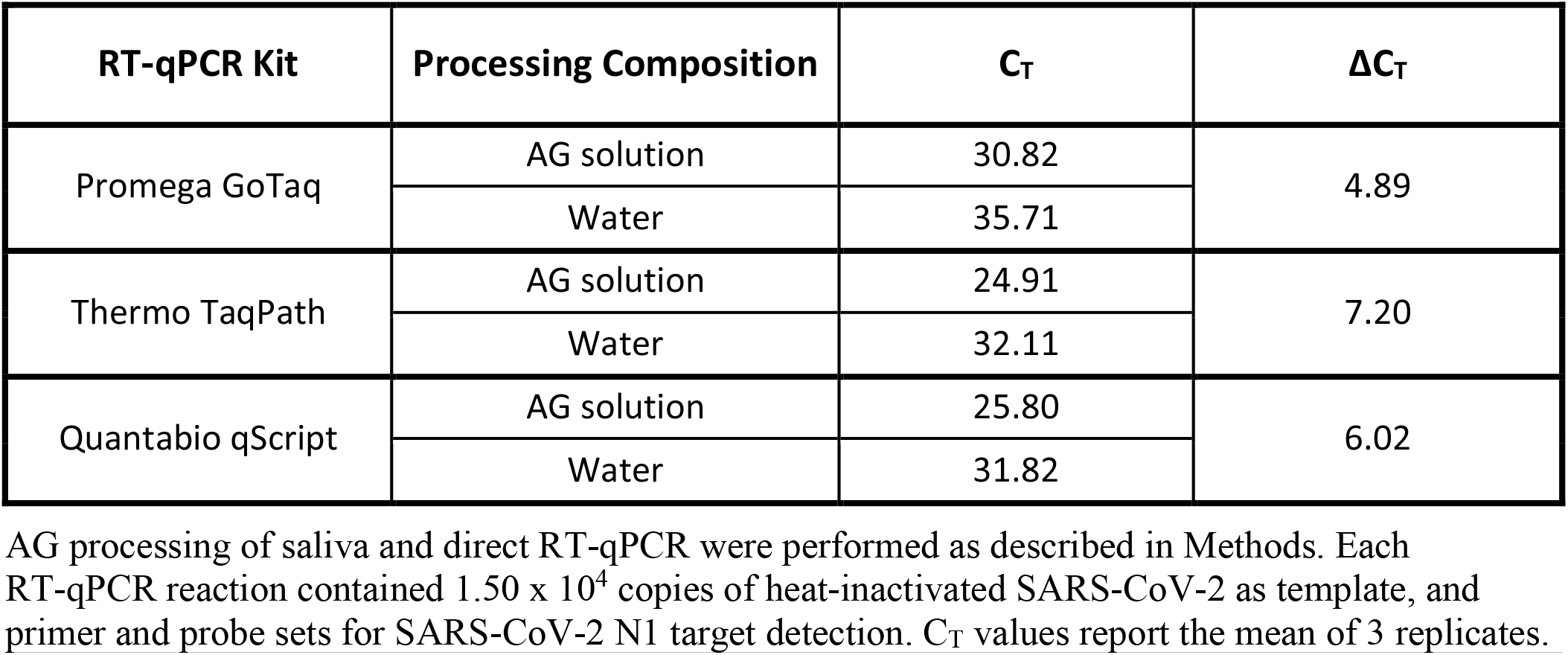
Detection of SARS-CoV-2 RNA in AG processed saliva using various RT-PCR kits.

| RT-qPCR Kit | Processing Composition | $C_T$ | $\Delta C_T$ |
| --- | --- | --- | --- |
| Promega GoTaq | AG solution | 30.82 | 4.89 |
|  | Water | 35.71 |  |
| Thermo TaqPath | AG solution | 24.91 | 7.20 |
|  | Water | 32.11 |  |
| Quantabio qScript | AG solution | 25.80 | 6.02 |
|  | Water | 31.82 |  |

AG processing of saliva and direct RT-qPCR were performed as described in Methods. Each RT-qPCR reaction contained $1.50 \times 10^4$ copies of heat-inactivated SARS-CoV-2 as template, and primer and probe sets for SARS-CoV-2 N1 target detection. $C_T$ values report the mean of 3 replicates.

AG processing can be combined with a heating step prior to RT-qPCR and/or the addition of a non-ionic detergent to the RT-qPCR assay. In our study, saliva containing inactivated virus was heated prior to AG processing and Tween-20 was added into the RT-qPCR reaction mix. **Table 4** shows the effects of heating and addition of Tween-20 on detection of SARS-CoV-2 RNA in the AG processed saliva. Heating saliva at 65 C or at 95 C substantially improved detection of viral RNA, as evidenced by a 5 C_T_ decrease in amplification cycles needed to detect SARS-CoV-2 RNA. Either heating or addition of Tween-20, when applied separately, have similar beneficial effects on detection of SARS-CoV-2 RNA; however, the combined effects of the two treatments were not additive. In the presence of Tween-20, heating was not necessary to achieve the optimal SARS-CoV-2 detection sensitivity reported in this study. However, heating clinical samples at > 56 C can provide an additional layer of safety by inactivating SARS-CoV-2 prior to handling [17]. In additional experiments, Tween-20 added directly to saliva samples instead of the RT-qPCR reaction mix caused a time-dependent degradation of SARS-CoV-2 RNA (results not shown).

**Table 4.** Effect of Tween-20 and heat on CT values for detecting SARS-CoV-2 in AG processed saliva.

| Treated<br>30 min<br>@ 95 C | Tween-20<br>0.2% | C<sub>T</sub> | Treated<br>30 min<br>@ 65 C | Tween-20<br>0.2% | C<sub>T</sub> |
| --- | --- | --- | --- | --- | --- |
| - | - | 30.09 | - | - | 31.81 |
| - | + | 26.31 | - | + | 24.44 |
| + | - | 24.92 | + | - | 26.26 |
| + | + | 24.94 | + | + | 23.95 |

Saliva specimens spiked with γ-irradiated SARS-CoV-2 were incubated for 30 min at room temperature, at 65 C, or at 95 C. After incubation, saliva specimens were AG processed and assayed by RT-qPCR using the KiCqStart One-Step RT-qPCR kit with the N1 primer and probe set. Each RT-qPCR reaction contained $1.00 \times 10^4$ copies of a viral template. As indicated, some RT-qPCR assays were supplemented with Tween®-20 (CAS 9005-64-5) to reach 0.2% concentration in the reaction mix. Each C<sub>T</sub> value reports the mean of 3 replicates.

AG processing of saliva with direct RT-qPCR allows a wide detection range (**Figure 1**). The detection range for γ-irradiated SARS-CoV-2 RNA in AG-processed saliva was from 400,000 copies to 4 copies per RT-qPCR reaction. C_T_ values ranged from 16.8 for 400,000 copies to 33.7 for 5 copies per reaction. Thus 400,000 and 4 copies per reaction correspond to x 10^8^ copies and 1.5 x 10^3^ copies of viral RNA per ml of saliva, respectively.

**Figure 1.**
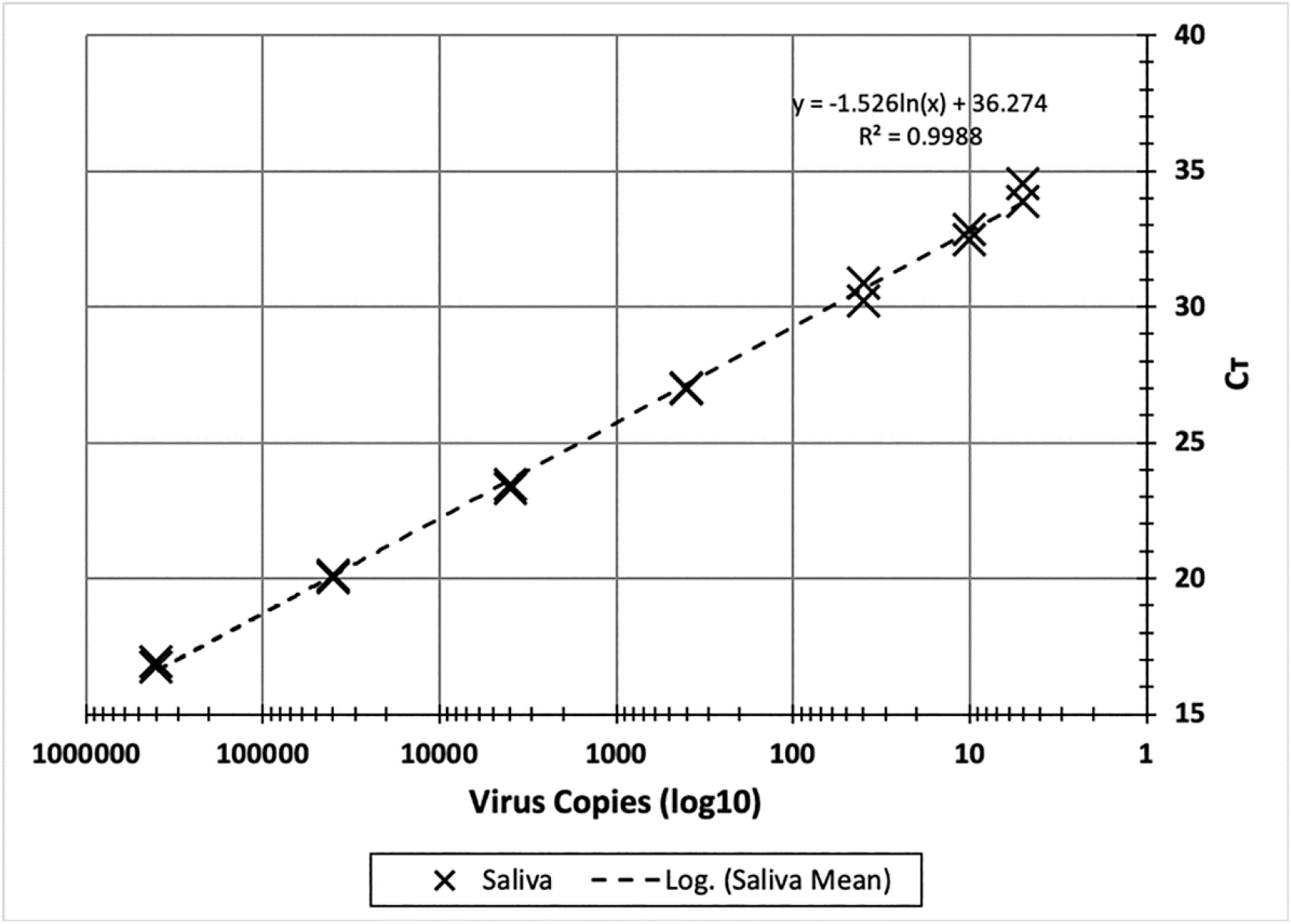
Detection range for SARS-CoV-2 in saliva using direct RT-qPCR with AG processing. AG processing solutions were prepared by mixing 2 volumes of saliva spiked with γ-irradiated SARS-CoV-2 and 1 volume of alkaline glycol solution. SARS-CoV-2 concentrations, per RT-qPCR reaction, ranged from 400,000 to 4 viruses. RT-qPCR was performed with 1 step KiCqStart RT-PCR kit with primer and probe set for N1 target detection. Shown are C_T_ values for each concentration in duplicate.

Human saliva collected from infected subjects is reported to contain from 10^10^ to 10^4^ SARS-CoV-2 virus particles per ml of saliva [5]. Thus, the method reported here – AG processing with direct RT-qPCR– provides an effective, sensitive, and robust diagnostic tool for detecting SARS-CoV-2.

It has been previously reported that saliva samples provide a more sensitive specimen for detecting SARS-CoV-2 in COVID-19 patients than nasopharyngeal swabs [5]. However, nasopharyngeal swabs are still a prevalent method of specimen collection for COVID-19 testing. Following collection, the oropharyngeal or nasopharyngeal swab specimens are immersed in UTM or Hanks ‘medium and used for COVID-19 testing. Specimen collection by swab does not provide a consistent amount of biological matter for diagnosis. In our tests of buccal swabs, the amount of swab-collected material varied over 2-fold, from 60 mg to 170 mg of specimen per swab. Nasopharyngeal swabs, being more difficult to collect, are likely to show even higher variability. Significant variability in the number of cells and viral particles in nasopharyngeal specimens was previously reported [18].

To simulate swab-derived clinical samples and provide a defined sample content for determining the detection range of viral RNA, we used compositions containing γ-irradiated SARS-CoV-2 in UTM or Hanks ‘medium and 20% (v/v) saliva. **Figure 2** shows that the detection range for SARS-CoV-2 RNA in UTM or Hanks ‘medium was similar to the viral detection range in AG-processed saliva. C_T_ values were 18.2 and 16.2 for 400,000 copies of SARS-CoV-2, and 35.5 and 32.2 for 5 copies of SARS-CoV-2 in UTM and Hanks ‘medium, respectively.

**Figure 2.**
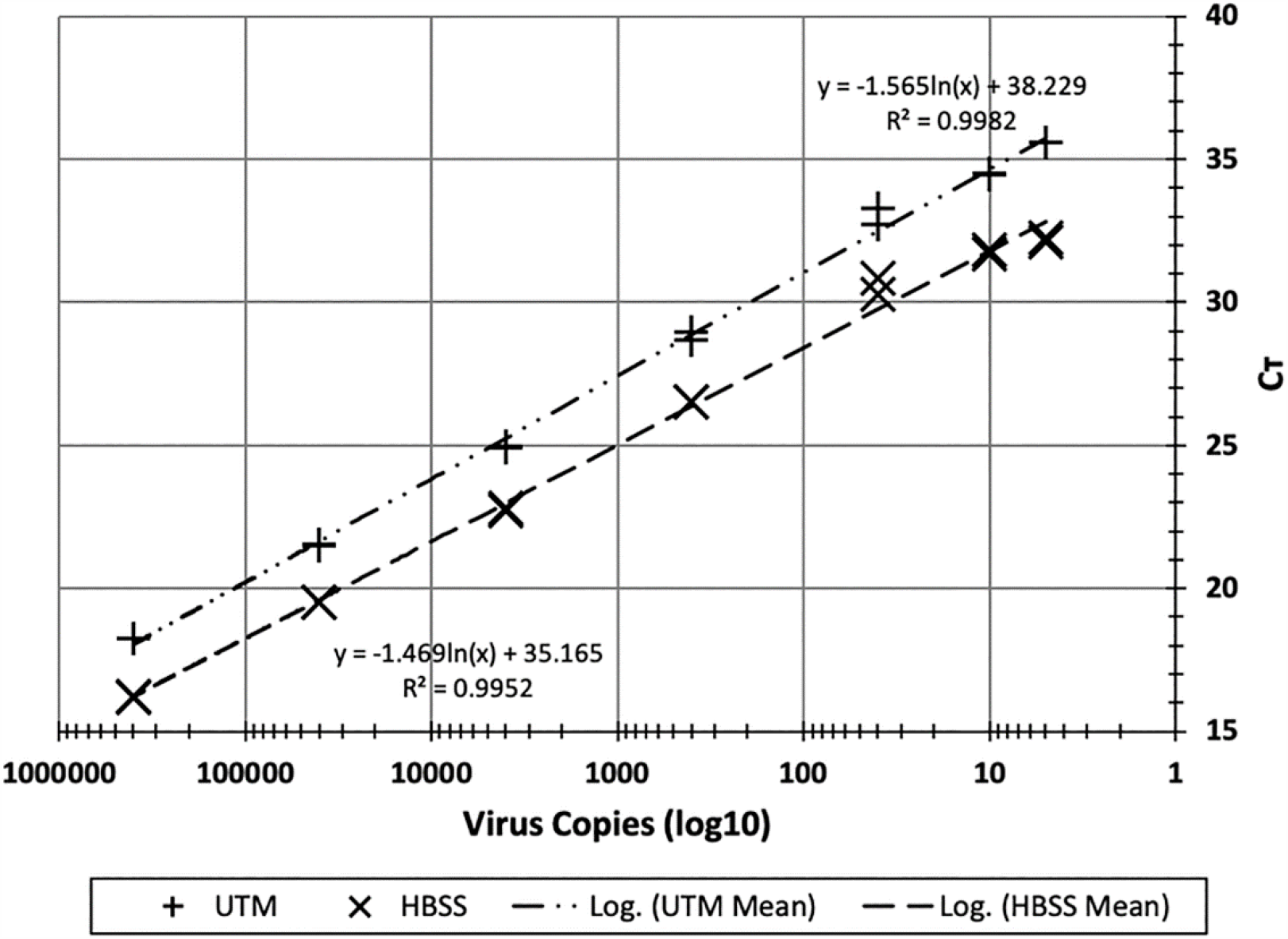
Detection range for SARS-CoV-2 in Universal Transport Medium for Viruses (UTM) or Hanks storage medium (HBSS) using AG processing with direct RT-qPCR. Samples were prepared by mixing 2 volumes of UTM or Hank’s containing 20% saliva and spiked with γ-irradiated SARS-CoV-2. For AG processing, 2 volumes of sample were mixed with 1 volume of alkaline glycol solution. The final SARS-CoV-2 concentrations ranged from 400,000 to 4 viruses per RT-qPCR reaction. RT-qPCR was performed with 1 step KiCqStart RT-PCR kit with primer and probe set for N1 target detection. Shown are CT for each concentration in duplicate.

To determine the lower limit of detection (LOD) for SARS-CoV-2 RNA copy number in AG-processed saliva, we performed RT-qPCR using the KiCqStart kit and saliva samples that contained single-digit numbers of virus particles per reaction (**Table 5**). Saliva was spiked with γ-irradiated SARS-CoV-2 virus to reach a final concentration of 3, 2, and 1 copy per reaction. Each viral concentration was tested in 20 replicates. In accord with the U.S. FDA regulation, determination of LOD requires 19 positive detections out of 20 repetitions, and the LOD is taken as twice the lowest mean value. Results in Table 5 show 20, 19, and 19 positive RT-qPCR results for 3, 2, and 1 copy per reaction, respectively. Using the FDA definition, the LOD for detecting copies per reaction, which corresponds to 606 viral copies per ml of saliva.

**Table 5.** Determination of LOD for detecting SARS-CoV-2 in RT-qPCR using AG processed saliva.

| SARS-CoV-2<br>copies per<br>reaction | Successful<br>amplification<br>(of 20) | C<sub>T</sub> | STDEV | Minimum<br>C<sub>T</sub> observed | Maximum<br>C<sub>T</sub> observed |
| --- | --- | --- | --- | --- | --- |
| 3 | 20 | 33.01 | 0.90 | 31.82 | 35.03 |
| 2 | 19 | 33.42 | 0.89 | 31.68 | 36.14 |
| 1 | 19 | 34.27 | 1.04 | 32.80 | 36.18 |

AG processing solutions were prepared by mixing two volumes of saliva spiked with γ-irradiated SARS-CoV-2 and one volume of alkaline glycol solution, to contain 3, 2, or 1 viral particle(s) per reaction. RT-qPCR was performed using the KiCqStart One-Step RT-qPCR kit with the N1 primer and probe set. Each C<sub>T</sub> value reports the mean of 3 replicates.

The detection sensitivity for samples treated with heat and/or Tween-20 and processed with the method is given in **Table 6**. In addition, detection sensitivity of the AG processing direct RT-qPCR assay can be increased an additional two-fold by increasing the RT-qPCR reaction volume from 20 µl to 50 µl (results not shown).

**Table 6.**
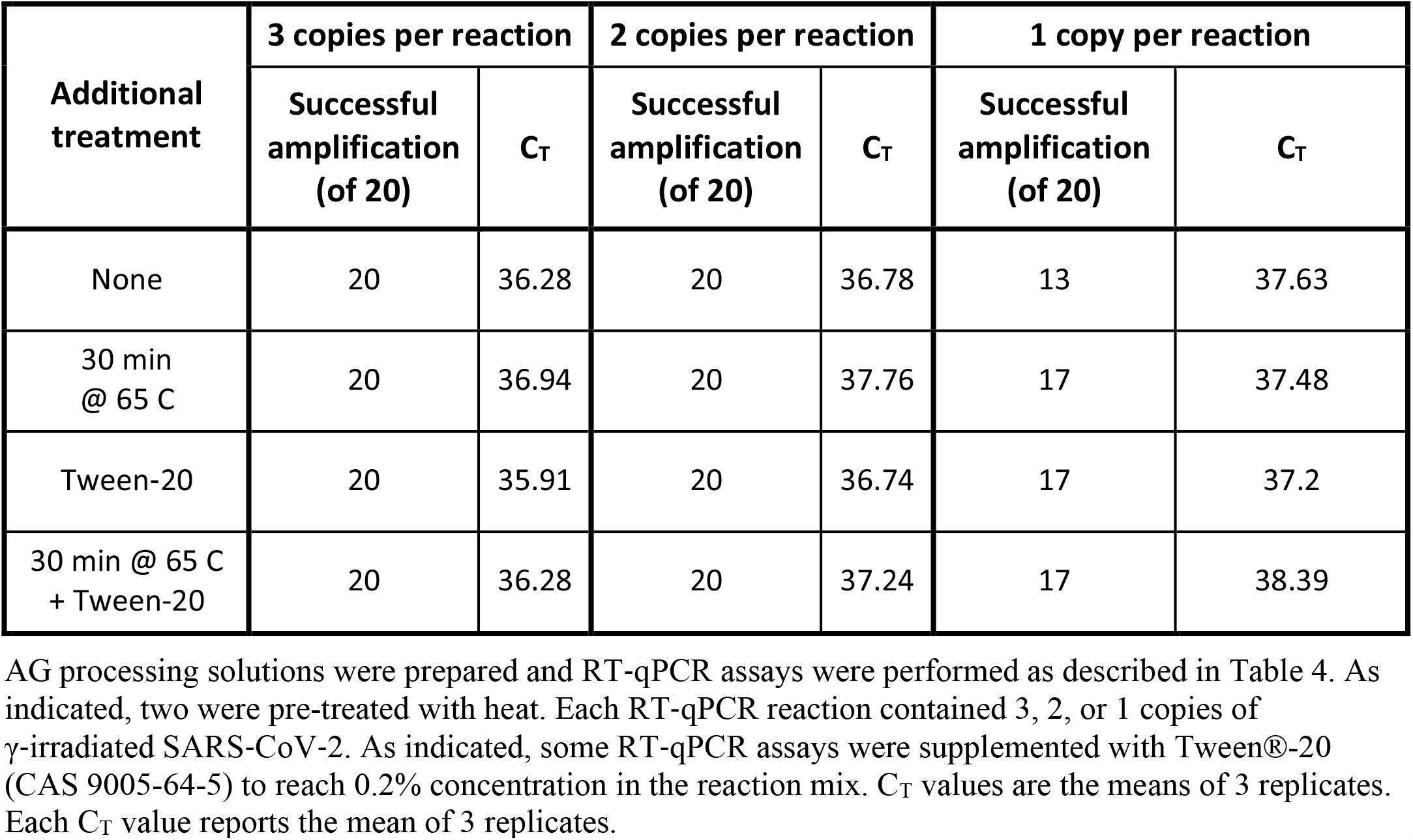
Effect of heating and Tween-20 on C_T_ using AG processed saliva with low viral copy number.

| Additional treatment | 3 copies per reaction |  | 2 copies per reaction |  | 1 copy per reaction |  |
| --- | --- | --- | --- | --- | --- | --- |
| | Successful amplification (of 20) | $C_T$ | Successful amplification (of 20) | $C_T$ | Successful amplification (of 20) | $C_T$ |
| None | 20 | 36.28 | 20 | 36.78 | 13 | 37.63 |
| 30 min @ 65 C | 20 | 36.94 | 20 | 37.76 | 17 | 37.48 |
| Tween-20 | 20 | 35.91 | 20 | 36.74 | 17 | 37.2 |
| 30 min @ 65 C + Tween-20 | 20 | 36.28 | 20 | 37.24 | 17 | 38.39 |

Additional validation of our method was provided by the diagnosis of COVID-19 in one sample donor. The donor provided a self-collected saliva specimen that was incubated at 95 C for 30 min then AG processed and used for detection of SARS-CoV-2 RNA by direct RT-qPCR with primers for the N1 target. Results of this test showed amplification of the target sequence with C_T_ 32.9 in the control samples without added reference material. In accordance with IRB requirements, the donor was notified of the results and instructed to consult a physician. The donor was diagnosed with COVID-19 following a positive result from an FDA-approved nasopharyngeal swab-based RT-PCR test.

## Conclusions

AG processing combined with direct RT-qPCR is an effective and highly sensitive method for detecting of SARS-CoV-2 without a requirement for viral RNA purification. The method has a sensitivity on par with or greater than existing methods that require an initial RNA extraction step. It is effective over a wide range of viral concentrations in either saliva specimens or samples collected by swab into media. AG processing is robust across different RT-qPCR platforms and works with multiple FDA-approved RT-qPCR products. These advantages, plus its simplicity and cost-effectiveness, can increase laboratory throughput and streamline COVID-19 diagnosis. Although this report focused on the use of AG processing for detection of SARS-CoV-2 in saliva or swab-derived oral specimens, the method can also be used for processing SARS-CoV-2 containing specimens derived from a variety of biological fluids and tissues. It is also easily adapted for processing specimens containing other RNA viruses.

## Data Availability

All dat presented in the manuscript are fully available

